# Improving safe vaginal deliveries using evidence-based practices at a semi-urban hospital in Dhaka, Bangladesh

**DOI:** 10.1101/2025.07.26.25332238

**Authors:** Anjuman Ara, Khurshid Talukder, Sanjida Akhter, Abdullah Asif, Saria Tasnim, Mohammad Khairul Islam, Soofia Khatoon, Ferdousi Begum

## Abstract

**Background:** Caesarean section (CS) rates in Bangladesh have risen dramatically, with some facilities reporting rates above 90%. Overuse of CS is associated with increased maternal and neonatal risks, underscoring the need for practical, evidence-based interventions to reduce unnecessary procedures. The objective of this study was to assess whether a package of evidence-based maternity practices, combined with routine monitoring, could reduce CS rates in a semi-urban hospital in Dhaka, Bangladesh.

**Methods:** We conducted a pre-post (two-period) evaluation using consecutive deliveries sampled in two independent time windows. The intervention was implemented at the Centre for Women and Child Health (renamed to Ashulia Women and Children Hospital as of 2022) between May 2017 and February 2019. Data were collected in two phases: baseline (n=1,116) and endline (n=1,252). A set of 11 practices was introduced to promote safe normal vaginal delivery, including antenatal counselling, improved labour monitoring, and promotion of vaginal birth after caesarean. Deliveries were classified according to the Robson Ten Group Classification System. Statistical analyses were performed using chi-squared tests.

**Results:** The overall CS rate declined from 52% at baseline to 42% at endline (p<0.001), representing a 19% relative reduction. Significant decreases were observed in Robson Groups 2a (p=0.017), 2b (p<0.001), 4a (p<0.001), 4b (p<0.001), and 5 (p=0.004). The intervention increased the proportion of women entering spontaneous labour (Groups 1 and 3) and reduced repeat CS through successful implementation of vaginal birth after caesarean. No adverse trends in maternal or neonatal outcomes were identified.

**Conclusion:** Implementation of a structured package of evidence-based obstetric practices, supported by systematic monitoring with the Robson classification, effectively reduced unnecessary CS in this hospital setting. These findings provide practical evidence for reducing CS rates while maintaining safety in similar low-resource contexts.

## Background

The caesarean section (CS) is one of the most commonly performed surgical interventions worldwide^1^, with significant implications for maternal and neonatal health. While essential in managing complications, its overuse has become a growing concern. The World Health Organization recommends that regional CS rates should remain between 10–15% to strike a balance between medical necessity and safety^2^. However, in the last three decades, CS rates have risen dramatically worldwide. Between 1990 and 2014, the global average increased from 6.7% to 19%, with an annual growth rate of 4.4%^3^. Regions such as Latin America and Asia have experienced the highest increases, reflecting changes in obstetric practices, healthcare systems, and societal preferences^3^.

Although surgical techniques and anaesthesia have advanced, unnecessary CS still poses significant risks compared to normal vaginal delivery (NVD)^4^. Studies indicate that unnecessary CS increases maternal morbidity, including infections, haemorrhage, and thromboembolism, along with long-term complications such as uterine rupture in subsequent pregnancies and greater incidence of post-partum depression^4–7^. NVD, on the other hand, remains safer and more cost-effective in uncomplicated deliveries, with median costs of NVD in Bangladesh estimated to be up to five times lower than CS procedures^8^. Distressingly, over one-third of the 18.5 million annual CS procedures conducted worldwide in 2008 were considered unnecessary, emphasizing the need for effective reduction strategies^9^.

Bangladesh exemplifies the complexities of rising CS rates in low- and middle-income countries. Over the past two decades, the national CS rate has rapidly escalated from 12% in 2010 to 31% in 2016^8^, and in 2023 reached a staggering rate of 51%^10^. Alarmingly, facility-based CS rates in Bangladesh have reached unprecedented levels, with private facilities reporting rates as high as 90%, compared to 55% in government facilities and 56% in NGO settings^10^. Urban women are twice as likely as their rural counterparts to deliver via CS (37% vs. 18%)^11,12^, and is most prevalent among women from wealthier, educated backgrounds or those attending frequent antenatal visits^10–12^. Projections suggest that, without targeted interventions, nearly all facility-based deliveries in the country could involve CS by 2030^13^.

Despite substantial advocacy from media^14,15^, professional bodies^16,17^, scientific groups^18,19^, and civil society^20,21^, active hospital-based efforts to reduce unnecessary CS remain largely underexplored in Bangladesh. A mix of factors, including the false perception of CS being safer^22^, patient preferences driven by family pressure and fear of labour pain^23^, and financial incentives for healthcare providers^22^, contribute to the high rates of CS in the country.

The Ashulia Women and Children Hospital (AWCH), a non-profit hospital based in Ashulia, Dhaka, manages approximately 1,500 deliveries annually. Between 2008 and 2016, AWCH – then known as Centre of Woman and Child Health (CWCH) – recorded CS rates ranging from 58% to 73% per year. Recognizing the need to address the high rates of CS, CWCH commenced a two-year intervention study in January 2017 to reduce CS rates, with the following objectives:

1. Increase safe NVDs through a package of 11 interventions implemented within AWCH’s existing maternity services.
2. Implement the Robson Ten Group Classification System (TGCS).
3. Monitor the monthly rate and indications of CS.

The package of 11 interventions was selected to address the multifactorial drivers of CS at AWCH. Strategies such as enhanced counselling and birth preparedness were intended to counteract women’s fear of labour pain and family pressure favouring CS. Consultant review of CS indications during ward rounds were introduced to reduce provider bias and financial incentive-driven decisions, and improved labour monitoring aimed to increase confidence in the safety of vaginal birth. Together, these measures were expected to create a supportive environment where normal vaginal delivery would be prioritised whenever medically appropriate.

This study evaluates the outcomes of the intervention programme, and its effectiveness in reducing CS rates through evidence-based practices and systematic classification using TGCS. Introduced in 2001^24^, the TGCS classifies deliveries into mutually exclusive categories based on parity, gestational age, labour onset, and foetal presentation, offering a practical tool for identifying and addressing drivers of CS^25^. Its adaptability has proven particularly valuable in resource-limited settings, enabling targeted interventions to address excessive CS rates^26,27^.

The design of the intervention package was guided by a combination of evidence-based obstetric practices and principles of quality improvement. The intervention design recognised that both provider practice and patient decision-making are shaped by incentives, perceptions of safety, and cultural expectations. By integrating structured classification using TGCS combined with targeted practice changes, it would enable a measurable and sustainable reduction in unnecessary CS.

Our intervention targeted the key proximal drivers of unnecessary caesarean section at CWCH: weak labour monitoring, low use of evidence-based induction/vaginal birth after caesarean (VBAC) criteria, limited consultant oversight of CS indications, and insufficient antenatal counselling. The 11-point package was designed to strengthen clinical decision-making and labour management so that avoidable CS are reduced without increasing adverse maternal or neonatal outcomes. We hypothesise the following causal pathway (Fig 1): improved competencies & documentation → better risk stratification and fewer inappropriate interventions → fewer unnecessary CS, while safety is maintained through consultant review and CTG/partograph monitoring.

**Fig 1.**
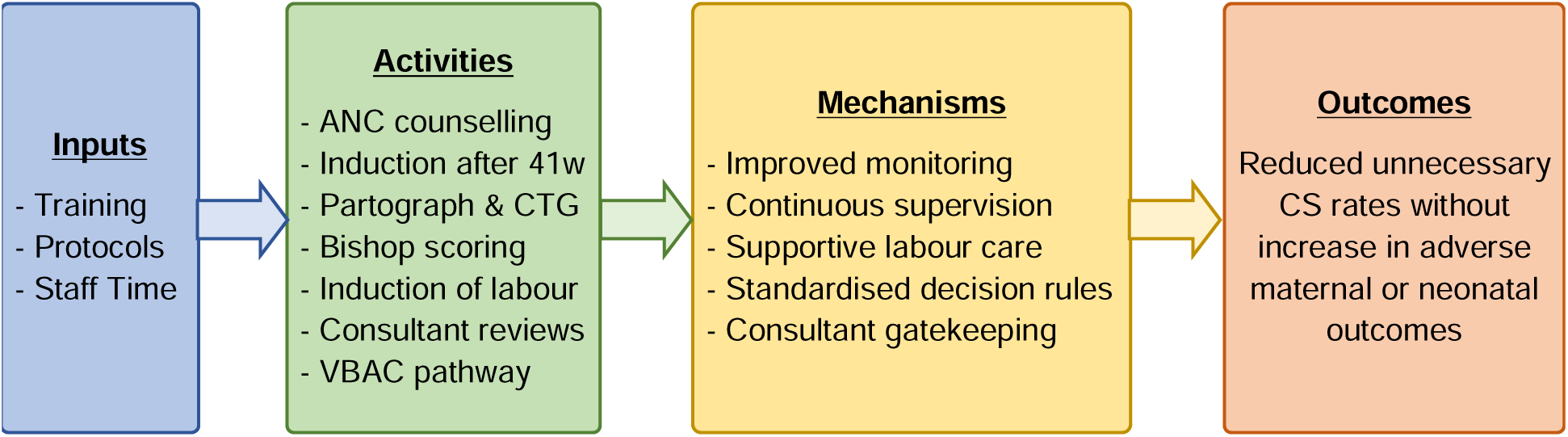
Logic model of the intervention package.

The findings of this study offer critical insights into the effectiveness of these interventions in reducing CS rates and their broader implications for addressing excessive CS rates in Bangladesh and similar settings. Given the rapid rise in CS rates and the limited hospital-based efforts to counter this trend, this study contributes towards understanding the complexities involved in reducing unnecessary CS by implementing evidence-based strategies.

## Methods

### Study context, design and setting

This was a hospital-based, pre-post intervention study using consecutive deliveries sampled in two independent time windows of a package of 11 maternity interventions designed to increase safe NVDs at AWCH.

At the time of the study, AWCH (then known as the Centre for Women and Child Health – CWCH) was a 100-bed, non-profit, single-storey, semi-urban hospital of 30,000 square feet situated in Ashulia, Dhaka. The hospital primarily served low- and middle-income women employed in the surrounding garment industry. It employed 388 full-time staff, including 98 doctors, 66 nurses, 44 patient care attendants (PCAs), and 180 support staff. The Obstetrics and Gynaecology (OBGYN) Department conducted approximately 1,500 deliveries annually. It had 30 inpatient beds, two labour rooms, and access to the hospital’s two operating theatres for CS. Staffing included a professor who served as Head of Department, nine consultants, 15 medical officers, 12 nurses, and eight PCAs.

Despite this infrastructure, CS rates were persistently high, influenced by provider preferences, family demand, and preexisting organisational norms. These contextual factors, combined with resource limitations and staff workload, were key considerations in designing the intervention.

### Intervention

A total of 15 medical officers and 12 nurses from the OBGYN Department were provided training on how to safely increase NVDs by AWCH’s Head of Department for OBGYN as well as nine other consultants. Three training sessions were conducted in total, all at the hospital premises. Topics that were covered through the three 10-hour interactive workshops on 7 to 8, 14 to 15, and 21 to 22 February 2018 included antenatal care (ANC), partograph use, cardiotocography (CTG) interpretation, data collection, TGCS classification and immediate newborn care. These training topics were subsequently discussed and reinforced through daily departmental morning sessions and ward rounds, which allowed for ongoing discussion, problem-solving, and supervision throughout the project period. This training was the initial rollout step within a broader implementation package that included protocol standardisation, supervision, audit-and-feedback, and routine TGCS-based monitoring.

The intervention package comprised 11 evidence-based maternity practices designed to reduce unnecessary CS and promote safe NVDs:

1. ANC counselling for expecting mothers.
2. Waiting up to 41 weeks of gestation for spontaneous onset of labour.
3. Risk screening for NVD and induction of labour at term.
4. Assessment of Bishop Score on admission.
5. Supportive care during labour and delivery.
6. Continuous monitoring with CTG.
7. Use of partograph
8. Induction or augmentation of labour as indicated, using prostaglandin or oxytocin for induction, based on Bishop Score.
9. Consultant review of CS indications during ward rounds.
10. Vaginal birth after caesarean (VBAC) for selected cases with appropriate monitoring.
11. Immediate care of the newborn.

A postgraduate OBGYN consultant acted as the Study Coordinator, overseeing routine data collection and quality assurance. Front-line consultants, medical officers, and nurses assigned Robson classifications at the time of delivery using semi-structured questionnaires, provided supportive care, recorded partograph and CTG findings, and assisted in newborn care. Fidelity to these practices was supported by consultant oversight, daily ward-round reviews, and systematic documentation on partographs, CTG records, and patient case notes. Elective CS required consultant approval and documented informed consent.

### Participants

The eligible study population included all women delivering at AWCH between 29 May 2017 and 10 February 2019, during Phase 1 and 2. Women who were admitted into the maternity and labour ward of AWCH were selected for this study. Consecutive sampling was used to select the participants. Women who had a stillbirth or declined consent to participate in the study were excluded.

### Data collection

Data collection was conducted in two rounds: baseline and endline. The data collected consisted of routine service data to monitor the mode of delivery and indications of CS. The project was divided into three phases – a baseline (Phase 1), an intervention, and an endline phase (Phase 2). We defined baseline as the pre-intervention period, from 01 June 2017 to 01 February 2018, approximately eight months, involved collecting data before and immediately after the intervention phase. The intervention phase consisted of providing training to healthcare workers from 7 to 22 February 2018. The training and initial rollout period was treated as a transition phase and excluded from the pre-post comparison. Endline data were collected during a later, post-implementation window in the last eight months of the study, from 11 June 2018 till 10 February 2019, to assess performance after routine practice stabilisation.

Routine delivery service data was collected at regular time intervals. This included the following variables: monthly data on the total number of deliveries, maternal age, the mode of delivery (CS or NVD), and the indication for CS if CS was done (absolute vs non-absolute indications). For TGCS classification, six basic obstetric variables were collected by maternity unit doctors on duty using a structured questionnaire, which included: gestational age in weeks, parity (nulliparous or multiparous), history of previous CS (yes or no), onset of labour (spontaneous, induced, pre-labour CS), number of foetuses (singleton or multiple), and foetal lie and presentation (cephalic presentation, breech presentation, or transverse lie).

### Data analysis

The primary outcomes were the monthly and annual CS rates, calculated from routine service data collected during the two data collection phases, and TGCS classifications. TGCS classifications were assigned to every single birth based on the six collected obstetric parameters. The TGCS classifies all deliveries into 12 mutually exclusive categories, outlined in Table 1.

**Table 1:** The Robson Ten Group Classification System.

| Robson TGCS Group | Description |
| --- | --- |
| 1 | Nulliparous, single cephalic, $\geq 37$ weeks, spontaneous labour |
| 2a | Nulliparous, single cephalic, $\geq 37$ weeks, induced labour |
| 2b | Nulliparous, single cephalic, $\geq 37$ weeks, CS before labour |
| 3 | Multiparous (without previous CS), single cephalic, $\geq 37$ weeks, spontaneous labour |
| 4a | Multiparous (without previous CS), single cephalic, $\geq 37$ weeks, induced labour |
| 4b | Multiparous (without previous CS), single cephalic, $\geq 37$ weeks, CS before labour |
| 5 | Multiparous, previous CS, single cephalic, $\geq 37$ weeks |
| 6 | Nulliparous with single breech |
| 7 | Multiparous with single breech (including previous CS) |
| 8 | All multiple pregnancies (including previous CS) |
| 9 | All abnormal lies (including previous CS) |
| 10 | All single cephalic, $< 37$ weeks (including previous CS) |

Secondary outcomes included CS indications which were divided into two categories of absolute and non-absolute indications, as defined by Stanton et al^28^. Absolute indications are situations where vaginal delivery is not possible or poses a highly significant risk to the mother or baby. Non-absolute indications are situations where a CS might be considered based on the potential risks and benefits for both the mother and the baby, and vaginal delivery may still be an option. In cases where mothers opted for a CS even after counselling was given, or where mothers declined to wait until 41 weeks of gestation, it was recorded as “maternal preference” under non-absolute indication.

Data was cleaned for any outliers and missing data before analysis. For entries that had missing variables, attempts were made to rectify by going through the raw data. If rectification was not possible, entries with partial or missing data/variables were dropped from the data analysis. Absolute reduction was calculated as (CS%_baseline_ - CS%_endline_) in percentage points. Relative reduction was calculated as (CS%_baseline_ - CS%_endline_) / CS%_baseline_ × 100.Pearson’s chi-squared tests were performed to calculate p-values in order to assess statistical significance for the parameters between the two phases. Fisher’s exact test was substituted in cases where the expected cell counts were less than five. For our study, a p-value less than 0.05 was considered statistically significant for a confidence interval of 95%. All analyses were conducted using SPSS version 18 (IBM Corp., Armond, New York)

### Ethics statement

Formal ethical approval for the study was obtained from the Ethical Review Committee of Centre of Woman and Child Health (approval number: CWCH/ERC/2017/010) on May 24, 2017. Written informed consent was obtained from all participants before data collection. Consent forms were in Bangla, and all willing participants provided their signature. For willing participants that could not read/write, the consent form was read out loud, and their thumb impression was obtained as proof of consent.

This manuscript was prepared in accordance with the Standards for Quality Improvement Reporting Excellence (SQUIRE) checklist to ensure comprehensive and transparent reporting^29^.

## Results

Prior to the intervention, service data from AWCH indicated that the incidence of CS remained consistently high between 2008 and 2016, ranging annually from 58% to 73% (Fig 2). The CS rate had declined to 55% among 1,577 deliveries at AWCH in 2017 during the baseline data collection phase, with a further reduction to 42% among 1,728 deliveries in 2018 after the implementation of the intervention package.

**Fig 2.**
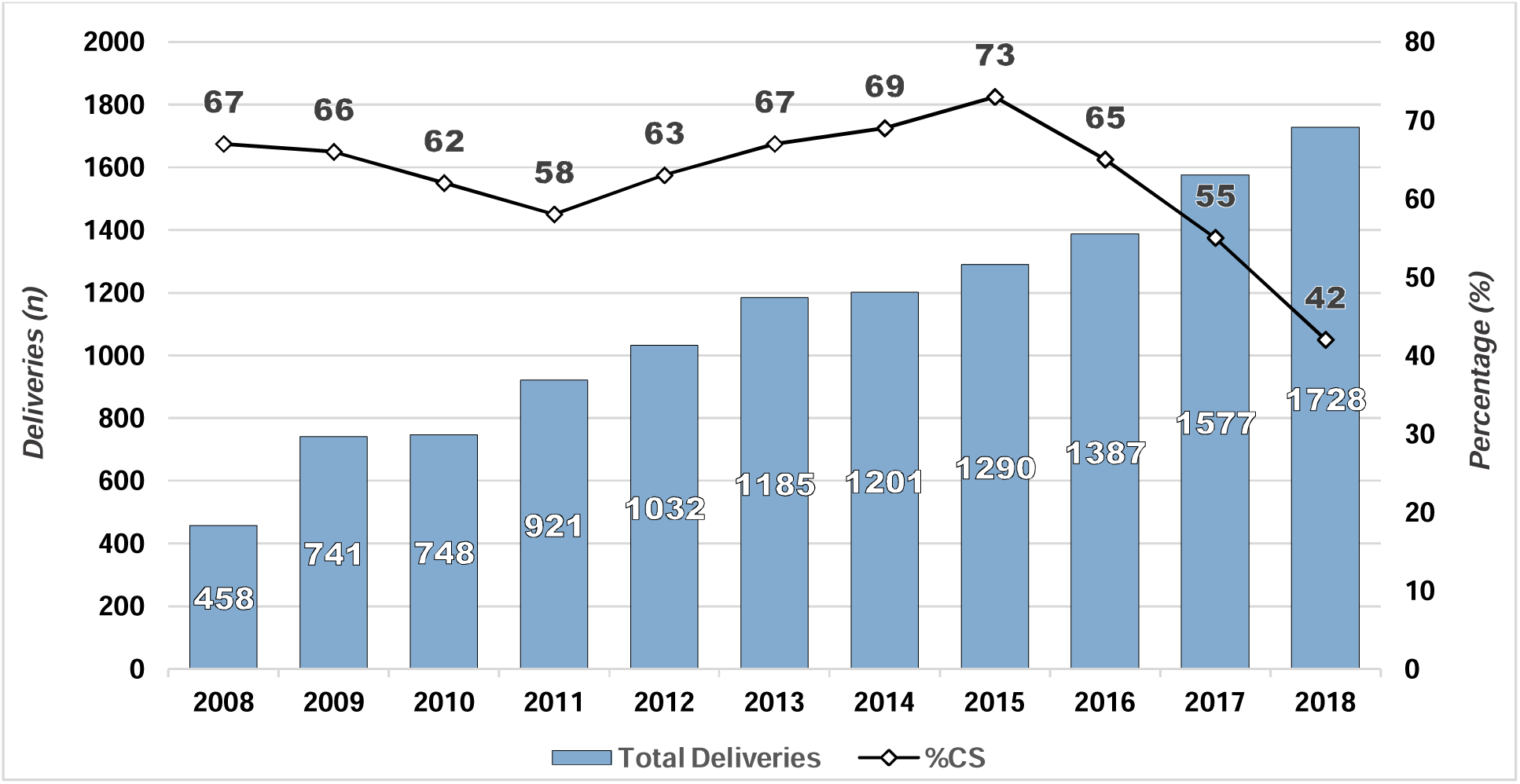
Yearly trends in total deliveries and CS rate at AWCH 2008-18.

A total of 1,198 women were screened for eligibility during Phase 1, of whom 1,176 met the inclusion criteria and were enrolled in the study. Following data validation and cleaning, 22 participants were excluded due to missing or wrong data, resulting in a final sample of 1,116 women for Phase 1. Similarly in Phase 2, 1,337 women were screened, and 1,299 were deemed eligible and enrolled. Data cleaning led to the exclusion of 38 participants, yielding a final sample of 1,252 women for Phase 2. A detailed comparison of CS incidence and indications is presented in Table 2. During Phase 1, 52% of women delivered by CS, compared to 42% in Phase 2, representing a statistically significant reduction (p <0.001). In both phases, approximately 5% of all CS were performed for absolute indications, with the remainder attributed to non-absolute indications as defined by Stanton et al^28^.

**Table 2.** Comparison of CS incidence and indications between Phase 1 and 2.

| Parameters | Phase 1 (N=1116) |  | Phase 2 (N=1252) |  | p-value |
| --- | --- | --- | --- | --- | --- |
|  | n | % | n | % |  |
| **Caesarean Deliveries** | **581** | **52.1** | **521** | **41.6** | **<0.001\*** |
| **Absolute indications for CS** | **29** | **5.0** | **34** | **6.5** | **0.273** |
| Obstructed Labour | 14 | 48.3 | 13 | 38.2 | 0.422 |
| Grade 3 or 4 placenta praevia | 4 | 13.8 | 2 | 5.9 | 0.401<sup>†</sup> |
|  | n | % | n | % |  |
| Impending uterine rupture | 3 | 10.3 | 12 | 35.3 | 0.036*† |
| Malpresentation (transverse, oblique, face) | 8 | 27.6 | 7 | 20.6 | 0.516 |
| **Non-absolute indications for CS** | **552** | **95** | **487** | **93.5** | **0.273** |
| Failure to progress in labour | 14 | 2.5 | 26 | 5.3 | 0.019* |
| Prolonged labour | 19 | 3.4 | 25 | 5.1 | 0.177 |
| Failed induction | 15 | 2.7 | 2 | 0.4 | 0.003*† |
| Previous caesarean delivery | 253 | 45.8 | 220 | 45.2 | 0.832 |
| Antepartum haemorrhage | 2 | 0.4 | 0 | 0 | - |
| Preeclampsia/eclampsia | 13 | 2.4 | 19 | 3.9 | 0.150 |
| Psychological indication | 2 | 0.4 | 0 | 0 | - |
| Maternal request | 33 | 6.0 | 24 | 4.9 | 0.458 |
| Precious (valuable) pregnancy | 24 | 4.3 | 17 | 3.5 | 0.479 |
| Foetal compromise including foetal distress, cord prolapse, severe IUGR | 152 | 27.5 | 125 | 25.7 | 0.497 |
| Breech presentation | 25 | 4.5 | 29 | 6.0 | 0.302 |

\*Statistically significant (p<0.05)
†Fisher's exact test applied due to small expected cell counts.
Note: Percentages in the main category rows are percent of all caesarean deliveries within that phase. Percentages in the subrows are percent of the main category. P-values for individual indications are exploratory and not adjusted for multiple comparisons; small cell sizes in some categories may limit statistical power.

Among absolute indications, a significant increase was noted in Phase 2 for impending uterine rupture (Phase 1: 10% vs Phase 2: 35%; p = 0.036). For non-absolute indications, significant differences were observed in the rates of CS for failure to progress in labour (2.5% vs 5.3%; p = 0.019) and failed induction (2.7% vs 0.4%; p = 0.003) between phases. In both phases, previous CS remained the most common indication (46% vs 46%; p = 0.958), followed by foetal compromise (28% vs 26%; p = 0.497).

The distribution of deliveries by the TGCS classification for both phases is provided in Table 3, following the World Health Organization’s Robson Implementation Manual^30^. In Phase 1, Group 5 (multiparous women with a previous CS, singleton, term, cephalic) comprised the largest proportion at 23%, followed by Group 3 (multiparous women without previous CS, singleton, term, cephalic, in spontaneous labour) at 22% and Group 1 (nulliparous women, singleton, term, cephalic, in spontaneous labour) at 16%. Group 5 also contributed the highest absolute and relative proportions to the overall CS rate at 23% and 44%, respectively. In Phase 2, Group 1 was the largest group at 32%, followed by Group 3 at 26%, with Group 5 falling to 20%. Similar to Phase 1, Group 5 continued to contribute the highest absolute and relative rates of CS, although the absolute rate declined to 18% while the relative contribution remained similar at 43%.

**Table 3:** Robson Classification for all deliveries in Phase 1 and 2.

| Robson TGCS Group | No. of women | No. of CS | Group Size<sup>a</sup> (%) | Group CS Rate<sup>b</sup> (%) | Absolute contribution to overall CS<sup>c</sup> (%) | Relative contribution to overall CS<sup>d</sup> (%) |
| --- | --- | --- | --- | --- | --- | --- |
| **Phase 1 Total** | **1116** | **581** | **100** | **52.1** | **52.1** | **100** |
| 1. Nulliparous, single cephalic, $\geq 37$ weeks, spontaneous labour | 180 | 34 | 16.1 | 18.9 | 3.0 | 5.9 |
| 2a. Nulliparous, single cephalic, $\geq 37$ weeks, induced labour | 91 | 46 | 8.2 | 50.5 | 4.1 | 7.9 |
| 2b. Nulliparous, single cephalic, $\geq 37$ weeks, CS before labour | 63 | 63 | 5.6 | 100 | 5.6 | 10.8 |
| 3. Multiparous (without previous CS), single cephalic, $\geq 37$ weeks, spontaneous labour | 241 | 26 | 21.6 | 10.8 | 2.3 | 4.5 |
| 4a. Multiparous (without previous CS), single cephalic, $\geq 37$ weeks, induced labour | 105 | 41 | 9.4 | 39.0 | 3.7 | 7.1 |
| 4b. Multiparous (without previous CS), single cephalic, $\geq 37$ weeks, CS before labour | 60 | 60 | 5.4 | 100 | 5.4 | 10.3 |
| 5. Multiparous, previous CS, single cephalic, $\geq 37$ weeks | 259 | 254 | 23.2 | 98.1 | 22.8 | 43.7 |
| 6. Nulliparous with single breech | 9 | 9 | 0.8 | 100 | 0.8 | 1.5 |
| 7. Multiparous with single breech (including previous CS) | 17 | 15 | 1.5 | 88.2 | 1.3 | 2.6 |
| 8. All multiple pregnancies (including previous CS) | 6 | 3 | 0.5 | 50.0 | 0.3 | 0.5 |
| 9. All abnormal lies (including previous CS) | 8 | 8 | 0.7 | 100 | 0.7 | 1.4 |
| 10. All single cephalic, $< 37$ weeks (including previous CS) | 77 | 22 | 6.9 | 28.6 | 2.0 | 3.8 |
| **Phase 2 Total** | **1252** | **521** | **100** | **41.6** | **41.6** | **100** |
| 1. Nulliparous, single cephalic, $\geq 37$ weeks, spontaneous labour | 394 | 107 | 31.5 | 27.2 | 8.5 | 20.5 |
| 2a. Nulliparous, single cephalic, $\geq 37$ weeks, induced labour | 51 | 30 | 4.1 | 58.8 | 2.4 | 5.8 |
| 2b. Nulliparous, single cephalic, $\geq 37$ weeks, CS before labour | 34 | 34 | 2.7 | 100 | 2.7 | 6.5 |
| 3. Multiparous (without previous CS), single cephalic, ≥37 weeks, spontaneous labour | 327 | 39 | 26.1 | 11.9 | 3.1 | 7.5 |
| 4a. Multiparous (without previous CS), single cephalic, ≥37 weeks, induced labour | 23 | 8 | 1.8 | 34.8 | 0.6 | 1.5 |
| 4b. Multiparous (without previous CS), single cephalic, ≥37 weeks, CS before labour | 19 | 19 | 1.5 | 100 | 1.5 | 3.6 |
| 5. Multiparous, previous CS, single cephalic, ≥37 weeks | 251 | 226 | 20.0 | 90.0 | 18.1 | 43.4 |
| 6. Nulliparous with single breech | 13 | 5 | 1.0 | 38.5 | 0.4 | 1.0 |
| 7. Multiparous with single breech (including previous CS) | 29 | 23 | 2.3 | 79.3 | 1.8 | 4.4 |
| 8. All multiple pregnancies (including previous CS) | 10 | 6 | 0.8 | 60.0 | 0.5 | 1.2 |
| 9. All abnormal lies (including previous CS) | 4 | 4 | 0.3 | 100 | 0.3 | 0.8 |
| 10. All single cephalic, <37 weeks (including previous CS) | 97 | 20 | 7.7 | 20.6 | 1.6 | 3.8 |

<sup>a</sup> no. of women in the group/total N women delivered in the hospital x 100
<sup>b</sup> no. of CS in the group/total N of women in the group x 100
<sup>c</sup> no. of CS in the group/total N of women delivered in the hospital x 100
<sup>d</sup> n of CS in the group/total N of CS in the hospital x 100

Two-tailed chi-square tests were performed to assess differences in CS rates between phases within each TGCS group (Table 4). Among non-breech, nulliparous women, significant differences were observed between phases in Group 1 (3.0% vs 8.5%; p < 0.001), where Phase 2 had a higher CS rate, and in Groups 2a (4.1% vs 2.4%; p = 0.017) and 2b (5.6% vs 2.7%; p < 0.001), where Phase 2 had lower CS rates than Phase 1. For multiparous women, CS rates were significantly lower in Phase 2 compared to Phase 1 in Group 4a (3.7% vs 0.6%; p < 0.001), Group 4b (5.4% vs 1.5%; p < 0.001), and Group 5 (22.8% vs 18.0%; p = 0.004). No significant differences were noted in the other remaining groups between the two phases.

**Table 4:**
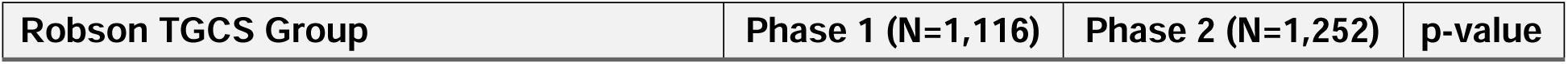
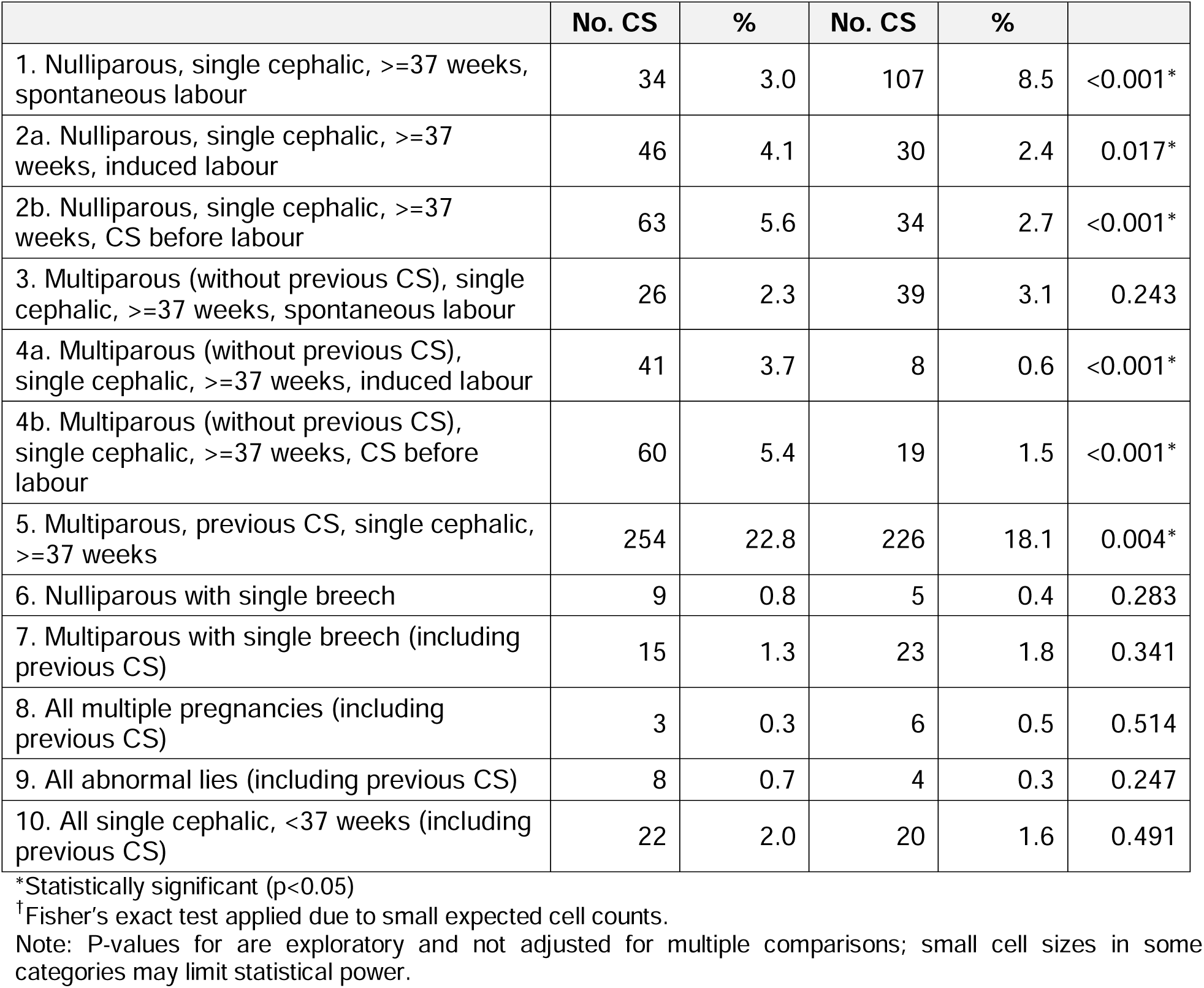
Comparison of absolute CS rate between Phase 1 and Phase 2 deliveries.

| Robson TGCS Group | Phase 1 (N=1,116) | Phase 2 (N=1,252) | p-value |
| --- | --- | --- | --- |

|  | No. CS | % | No. CS | % |  |
| --- | --- | --- | --- | --- | --- |
| 1. Nulliparous, single cephalic, $\geq 37$ weeks, spontaneous labour | 34 | 3.0 | 107 | 8.5 | $<0.001^*$ |
| 2a. Nulliparous, single cephalic, $\geq 37$ weeks, induced labour | 46 | 4.1 | 30 | 2.4 | 0.017* |
| 2b. Nulliparous, single cephalic, $\geq 37$ weeks, CS before labour | 63 | 5.6 | 34 | 2.7 | $<0.001^*$ |
| 3. Multiparous (without previous CS), single cephalic, $\geq 37$ weeks, spontaneous labour | 26 | 2.3 | 39 | 3.1 | 0.243 |
| 4a. Multiparous (without previous CS), single cephalic, $\geq 37$ weeks, induced labour | 41 | 3.7 | 8 | 0.6 | $<0.001^*$ |
| 4b. Multiparous (without previous CS), single cephalic, $\geq 37$ weeks, CS before labour | 60 | 5.4 | 19 | 1.5 | $<0.001^*$ |
| 5. Multiparous, previous CS, single cephalic, $\geq 37$ weeks | 254 | 22.8 | 226 | 18.1 | 0.004* |
| 6. Nulliparous with single breech | 9 | 0.8 | 5 | 0.4 | 0.283 |
| 7. Multiparous with single breech (including previous CS) | 15 | 1.3 | 23 | 1.8 | 0.341 |
| 8. All multiple pregnancies (including previous CS) | 3 | 0.3 | 6 | 0.5 | 0.514 |
| 9. All abnormal lies (including previous CS) | 8 | 0.7 | 4 | 0.3 | 0.247 |
| 10. All single cephalic, $< 37$ weeks (including previous CS) | 22 | 2.0 | 20 | 1.6 | 0.491 |

\*Statistically significant ( $p < 0.05$ )
†Fisher's exact test applied due to small expected cell counts.
Note: P-values for are exploratory and not adjusted for multiple comparisons; small cell sizes in some categories may limit statistical power.

## Discussion

There have been many reports in the media related to dangerous increases in rates of unnecessary caesarean sections throughout the country over the past decade. In addition, stakeholder and professional body discussions have taken place about these increases and strategies to decrease them. In 2019, a Public Interest Litigation petition was filed, and the High Court issued a rule upon the Ministry of Health and Family Welfare, Directorate General of Health Services, and Bangladesh Medical and Dental Council to explain why they had not taken action to stop unnecessary C-sections in hospitals and clinics, and why this failure should not be considered illegal^16^. In November 2019, the Directorate General of Health Services responded to the writ by issuing comprehensive national guidelines to promote normal delivery and curb unnecessary surgery^31^. In October 2023, a lag in implementation prompted the High Court to issue a directive requiring execution of the 2019 National Guidelines within six months^32^. Unfortunately, there has been no further action on implementation of these government guidelines since then.

To our knowledge, this study represents the first systematic effort to reduce facility-based CS rates in Bangladesh. This study demonstrates that a multifaceted intervention strategy at this hospital was effective in reducing the overall CS rate by 19% relatively, from 52% in baseline to 42% in endline. In comparison, latest available data indicates national CS averages of 90% in private health facilities and 56% in NGO-run hospitals^10^. This reduction aligns with findings from intervention studies conducted in other countries to reduce CS rates, where reductions have ranged from 34% in a Slovakian study to 12% in a Brazilian study^33,34^.

The use of the TGCS was particularly valuable in our study for systematically monitoring caesarean section trends across defined obstetric groups. A similar study from India utilizing an intervention package reported a significant reduction in the CS rate over a span of seven years, decreasing from 52% to 18%^35^. In this study, the TGCS was utilized for identifying groups that needed targeted tailored interventions for reducing CS. The TGCS provides valuable data for groups such as multiparous women with no previous CS undergoing a CS, large numbers of nulliparous women having CS before labour, and large numbers of multiparous women having CS due to previous uterine scar, allowing us to design targeted interventions for specific groups.

A principal component of the intervention package was antenatal counselling regarding the benefits and safety of NVD, which proved instrumental in encouraging both nulliparous and multiparous women without a previous CS (i.e. TGCS Groups 2b and 4b) to await spontaneous onset of labour up to 41 weeks rather than opting for early elective CS. As a result, the number of women classified within Groups 1 and 3 increased substantially in Phase 2 compared to Phase 1, reflecting a transfer from Groups 2b and 4b to Groups 1 and 3, respectively. Similarly, women who would otherwise have undergone early induction and been categorised in Groups 2a and 4a were encouraged to await spontaneous labour, contributing further to the increased number of women in Groups 1 and 3 in Phase 2.

However, this approach increased the CS rate within Group 1, a difference that was statistically significant, with a modest but statistically insignificant increase in Group 3. This increase is attributed to a subset of women who, despite awaiting spontaneous labour up to 41 weeks, failed to progress to delivery and required CS, often after failed induction or without the opportunity for timely induction. This is supported by the significant rise in CS indications for “Failure to progress in labour” in Phase 2 compared to Phase 1 (Table 2). Despite CS rate increases within Groups 1 and 3, the overall CS rate decreased across almost every other TGCS group, contributing to the net reduction in CS rates across the hospital.

Additionally, significant reductions in CS rates were observed within Groups 2a and 4a (nulliparous and multiparous women undergoing induction), underscoring the effectiveness of improved induction and labour management practices introduced during the intervention. These practices included induction at term, systematic monitoring using partograph and CTG, assessment of cervical readiness using the Bishop score, and the use of prostaglandins or oxytocin for induction based on the Bishop score. Such measures increased the likelihood of successful NVD following induction, thereby reducing unnecessary CS in these groups.

Importantly, the intervention also led to a significant reduction in the CS rate within Group 5 (multiparous women with a previous CS). This outcome highlights the successful implementation of strategies to promote VBAC in selected cases. Consultant-led reviews of CS indications during ward rounds, coupled with close intrapartum monitoring, facilitated the safe conduct of VBAC, contributing to a reduction in repeat CS while maintaining maternal and neonatal safety.

The use of TGCS throughout this study provided a structured and transparent method for monitoring CS trends, enabling the identification of groups where reductions were most effective and where additional interventions may be required. The observed reduction in overall CS rates, particularly in groups where CS is often performed for non-absolute indications, suggests that a judicious, data-driven approach can effectively address unnecessary CS while ensuring safety in facility-based deliveries.

Overall, these findings suggest that a targeted, context-specific intervention package incorporating antenatal counselling, evidence-based induction practices, and promotion of VBAC can effectively reduce unnecessary caesarean deliveries while maintaining the safety of mothers and newborns. At the end of this study, sustainability steps were embedded in routine practice and are still continuing as of December 2025: monthly TGCS reports are generated by a trained OBGYN registrar and reviewed during routine departmental meetings, senior consultants provide ongoing supervision and gatekeeping for CS decisions, newly hired OBGYN staff undertake a mandatory short training on the package and are supervised during an induction period. These institutionalised processes are the mechanisms we expect to maintain effect beyond the study period.

## Limitations

This study has certain limitations that should be acknowledged. First, as this study was conducted in a single semi-urban, non-profit hospital, the external generalisability of the results is limited. Facilities with different case-mixes, resource availability or provider incentives may not achieve similar reductions in CS rates. Second, the pre-post design lacked a concurrent control group. Without randomisation or parallel comparison, we cannot fully exclude secular trends or other external influences that may have contributed to the observed decline in CS rates. In addition, the relatively short follow-up may not capture longer-term sustainability of practice changes. Third, the multifaceted intervention obscures identification of the individual contribution of each component. The absence of prospectively collected quantitative fidelity metrics (for example, % of deliveries with completed partograph or documented consultant review) limits our ability to attribute the observed reduction in CS to any specific component of the package or to quantify adherence. Finally, patient-level factors such as women’s preferences, socio-economic status and background were not formally assessed. Future studies should consider incorporating mixed-methods approaches and multi-centre designs to validate and extend these findings.

## Conclusion

This intervention study conducted at AWCH demonstrates that a targeted, evidence-based intervention package can effectively reduce unnecessary CS within a facility-based setting in Bangladesh. The implementation of these interventions not only promoted NVDs but also ensured maternal and neonatal safety, particularly by supporting VBAC in selected cases. Given the global and national concerns surrounding the overuse of CS, this study contributes valuable insights into practical, scalable strategies for promoting safe, facility-based deliveries that can be adopted by other hospitals in their efforts to reduce CS to a medically justifiable rate.

## Data Availability

All data produced are available online at OPENICPSR with project ID 236841

https://www.openicpsr.org/openicpsr/project/236841

## Declarations

### Consent for publication

Not applicable

### Availability of data and materials

The data that support the findings of this study are available in a public, open access repository at OPENICPSR repository, https://doi.org/10.3886/E236841V536.

### Author Contributions

Anjuman Ara and Khurshid Talukder: conceptualization, investigation, methodology, project administration, and writing (original draft). Sanjida Akhter: conceptualization, investigation, supervision, and writing (review & editing). Abdullah Asif: data curation, formal analysis, validation, and writing (original draft). Saria Tasnim: methodology, supervision, and writing (review & editing). Mohammad Khairul Islam: data curation, investigation, and writing (review & editing). Soofia Khatoon: methodology and writing (review & editing). Ferdousi Begum: investigation and writing (review & editing).

## Acknowledgements

The authors would like to express their sincere gratitude to DrDabirUddinAhmed, CEO of AWCH, for his unwavering support and guidance throughout the planning and implementation of this study. We are deeply grateful to all the maternity ward and labour room doctors, nurses, and midwives for their dedication in delivering high-quality care, for embracing the new interventions, and their commitment to meticulous data collection. Special thanks are due to the customer care and administrative staff for their assistance in scheduling training sessions and for ensuring smooth logistics throughout the study period. Finally, we extend our gratitude to the women who participated in this study. Without their willingness and trust, this work would not have been possible. Their cooperation and feedback have been instrumental in shaping the interventions and improving our maternity services.

